# High excess mortality during the COVID-19 outbreak in Stockholm Region areas with young and socially vulnerable populations

**DOI:** 10.1101/2020.07.07.20147983

**Authors:** Amaia Calderón-Larrañaga, Davide L Vetrano, Debora Rizzuto, Tom Bellander, Laura Fratiglioni, Serhiy Dekhtyar

## Abstract

**Background:** We aimed to describe the distribution of excess mortality (EM) during the first weeks of the COVID-19 outbreak in the Stockholm Region, Sweden, according to individual age and sex, and the sociodemographic context

**Methods:** Weekly all-cause mortality data were obtained from Statistics Sweden for the period 01/01/2015 to 17/05/2020. EM during the first 20 weeks of 2020 was estimated by comparing observed mortality rates with expected mortality rates during the five previous years (N=2,379,792). EM variation by socioeconomic status (tertiles of income, education, Swedish-born, gainful employment) and age distribution (share of 70+ year-old persons) was explored based on Demographic Statistics Area (DeSO) data.

**Findings:** An EM was first detected during the week of March 23-29 2020. During the peaking week of the epidemic (6-12 April 2020), an EM of 160% was observed: 211% in 80+ year-old women; 179% in 80+ year-old men. During the same week, the highest EM was observed for DeSOs with lowest income (171%), lowest education (162%), lowest share of Swedish-born (178%), and lowest share of gainfully employed (174%). There was a 1.2 to 1.7-fold increase in EM between those areas with a higher vs. lower proportion of young people.

**Interpretation:** Living in areas with lower socioeconomic status and younger populations is linked to COVID-19 EM. These conditions might have facilitated the viral spread. Our findings add to the well-known biological vulnerability linked to increasing age, the relevance of the sociodemographic context when estimating the individual risk to COVID-19.

**Funding:** None.

## Introduction

In just three months since the first death attributed to COVID-19 was reported in Sweden on March 11 2020, the pandemic has claimed more lives than breast cancer and prostate cancer combined over an entire year in 2017.^1^ Although the high mortality due to COVID-19 is hardly disputed, current estimates of deaths may be underestimated as figures based on laboratory-confirmed results miss the false negative cases and those who were not tested at all. Further, they do not include the deaths caused by conditions that would have normally been treated, had hospitals not been overwhelmed by a surge of patients needing intensive care. In areas with extensive testing also overestimation is possible, since deaths in persons that tested positive may be unrelated to COVID-19. Excess mortality, the gap between the deaths from any cause, and the historical average for the same place and time of year, offers a more comprehensive way to measure the mortality linked to the COVID-19 outbreak. Assessing excess deaths in Sweden is especially relevant, as restrictions and confinement have been considerably less widespread compared with the rest of Europe.

It is known that older adults bear a disproportionate burden of COVID-19 mortality, with 89% of all deaths due to coronavirus as of June 22, 2020 in Sweden occurring in individuals aged 70 or above.^2^ Moreover, COVID-19 appears to impact socioeconomically vulnerable populations especially hard. A preliminary analysis of excess mortality between 1-10 April 2020 in the Stockholm Region (the area most affected by COVID-19 in Sweden), has revealed that excess deaths were highest in municipalities with lower education, income, and share of Swedish-born residents (*manuscript under review*). Few attempts have been made to integrate biological (old age) and social (economic deprivation) vulnerabilities in order to understand the force of mortality associated with the COVID-19 pandemic in Sweden. A recent study has found that in older adults, individual measures of socioeconomic deprivation, such as reduced disposable income or lower educational attainment appeared less predictive of COVID-19 mortality than in the working-age subset of the Swedish population.^3^

Individual-level influences of socioeconomic factors represent only one dimension of vulnerability faced by the older adults. Contextual socioeconomic characteristics have been described as another contributor to social disparities in older adults’ health,^4^ and are likely involved in the shaping of COVID-19 outcomes as well. Furthermore, the consequences of contextual deprivation are likely non-uniform depending on the predisposing conditions that may either accentuate or lessen the effect of individual socioeconomic vulnerabilities. Contextual demography may be one such factor. Preliminary data from New York City has shown that areas with a higher share of the population under the age of 18 had more COVID-19 cases, although it was unclear whether children accelerated the transmission, or whether reduced incomes underpinned this association.^5^ A recently proposed segmentation and shielding strategy places emphasis not only on those most vulnerable to COVID-19 outcomes, but also on their closest contacts and networks, who can transmit the disease to them.^6^ Therefore, it is important to assess how contextual age distribution and socioeconomic status interact in shaping COVID-19 excess deaths.

Our aim in this study is threefold: 1) To estimate the age- and sex-specific excess mortality during the first weeks of the COVID-19 outbreak in the Stockholm Region; 2) To explore to what extent COVID-19 excess mortality varies among socioeconomically different regional areas; and 3) To assess if the excess mortality variation linked to socioeconomic characteristics is modified by the age distribution of the population in the area.

## Methods

Weekly all-cause mortality figures and population data for the Stockholm region were provided by Statistics Sweden (SCB) for the period 1 January 2015 to 31 May 2020. Data from the two last weeks in 2020 (i.e. 18-31 May) were discarded due to quality considerations since there is a lag in data reporting by the Swedish Tax Agency to SCB. Both total and age- and sex-specific mortality rates were calculated for each week. The size of the resident population averaged across 13 weeks (3 months) was used as denominator for each trimester. At the beginning of 2020, 2,379,792 persons were resident in the Stockholm Region. The expected weekly mortality rates for the first 20 weeks of 2020 were obtained by averaging weekly mortality estimates for the years 2015 to 2019. Observed weekly mortality rates were compared with these in order to obtain excess mortality estimates: ((*observed rate - expected rate*) */ expected rate*) *\* 100*.

Mortality rates by socioeconomic (i.e. income, level of education, share of Swedish-born, share gainfully employed) and demographic (i.e. share of 70+ year-old persons) indicators for Demographic Statistics Areas (DeSO) within the Stockholm Region were also obtained from SCB. DeSOs are contiguous areas with around 1500 inhabitants (range: 700 and 2700) aiming at small within-area and large between-area socioeconomic variability. DeSO subdivisions are based on the boundaries of municipalities, election districts and major over 1000 inhabitants, and are considered to be stable over time.^7^

Income was measured as median employment and business (acquisition) income, and level of education as the share of above elementary (12 years) education across DeSOs. A person that is gainfully employed should have worked for at least one hour per week in the month of November, including those temporarily absent. If the administrative records do not contain information on working hours, income information is used by SCB instead. Data on DeSO-level sociodemographics corresponded to years 2018-2019. Socioeconomic indicators were categorized into low, medium or high according to tertiles of the DeSO distribution, and further divided according to the share of 70+ year-old persons (below vs. above the median) within each tertile.

Stata 16 (StataCorp) has been used for all the analyses. No ethical approval was needed for the present study given the use of aggregate-level data.

## Results

From 11 March 2020 – date of the first death attributed to COVID-19 – to 17 May 2020, 5234 people died in the Stockholm Region, compared to an average of 3277 during the same period in 2015-2019, i.e. an estimated excess of 1957 deaths. Starting in week 9-15 2020, all-cause mortality exhibited an ascending trend, peaking during the week 6-12 April 2020, and a subsequent decline. During the ascending phase, mortality rate increased from 12.7 to 33.0/100,000 inhabitants (**Figure 1**). It should be noted that no corresponding acceleration of mortality was observed for the same weeks in the previous five years; on the contrary, there was a trend of decreasing mortality in the corresponding period. During the week 6-12 April 2020, overall mortality exceeded by 160% (130% for women, 176% for men) that observed on average in the same week in the preceding five years. The excess varied across age and sex. In fact, in the same week, mortality rates were increased by 69% (49% for women and 82% for men) in individuals 0-64 years old, by 129% (68% for women and 175% for men) in individuals 65-79 years old, and by 193% (210% for women and 179% for men) in individuals 80 years old or older (**Table 1**).

**Table 1.** All-cause mortality rates (per 100,000 persons) and excess mortality in the Stockholm Region (N=2,379,792), Sweden, by age group and sex during the COVID-19 outbreak (i.e. March 9^th^ to April 12^th^ 2020).

| **Women** |  |  |  |  |  |  |  |  |  |
| --- | --- | --- | --- | --- | --- | --- | --- | --- | --- |
| **Week** | **<65 years** |  |  | **65-79 years** |  |  | **80+** |  |  |
|  | **Observed rate** | **Expected rate** | **Excess (%)** | **Observed rate** | **Expected rate** | **Excess (%)** | **Observed rate** | **Expected rate** | **Excess (%)** |
| **Mar 9-15** | 2.3 | 2.0 | 14.9 | 18.1 | 26.4 | -31.5 | 284.8 | 242.9 | 17.3 |
| **Mar 16-22** | 1.5 | 1.5 | 1.5 | 25.9 | 27.4 | -5.5 | 244.5 | 237.7 | 2.9 |
| **Mar 23-29** | 2.0 | 1.7 | 17.0 | 30.4 | 29.7 | 2.4 | 420.5 | 238.6 | 76.2 |
| **Mar 30-Apr 5** | 1.8 | 1.7 | 0.2 | 53.6 | 25.6 | 109.9 | 538.8 | 257.5 | 109.3 |
| **Apr 6-12** | 2.5 | 1.7 | 48.8 | 47.8 | 28.6 | 67.5 | 682.2 | 219.6 | 210.6 |
| **Men** |  |  |  |  |  |  |  |  |  |
| **Week** | **<65 years** |  |  | **65-79 years** |  |  | **80+** |  |  |
|  | **Observed rate** | **Expected rate** | **Excess (%)** | **Observed rate** | **Expected rate** | **Excess (%)** | **Observed rate** | **Expected rate** | **Excess (%)** |
| **Mar 9-15** | 2.6 | 2.7 | -3.0 | 33.4 | 35.9 | -7.0 | 216.0 | 307.4 | -29.8 |
| **Mar 16-22** | 2.6 | 2.7 | -5.9 | 42.7 | 38.2 | 11.8 | 372.0 | 265.3 | 40.2 |
| **Mar 23-29** | 2.8 | 3.0 | -6.5 | 61.9 | 37.7 | 64.0 | 482.7 | 264.6 | 82.4 |
| **Mar 30-Apr 5** | 5.0 | 2.8 | 77.7 | 82.5 | 38.0 | 117.4 | 609.0 | 286.4 | 112.6 |
| **Apr 6-12** | 5.0 | 2.8 | 81.6 | 103.8 | 37.8 | 174.7 | 757.0 | 271.0 | 179.4 |

Note: Excess mortality calculated comparing the weekly mortality rates during the weeks 11-15 of 2020 with the average mortality rate recorded for the corresponding weeks during the five previous years.

**Figure 1.**
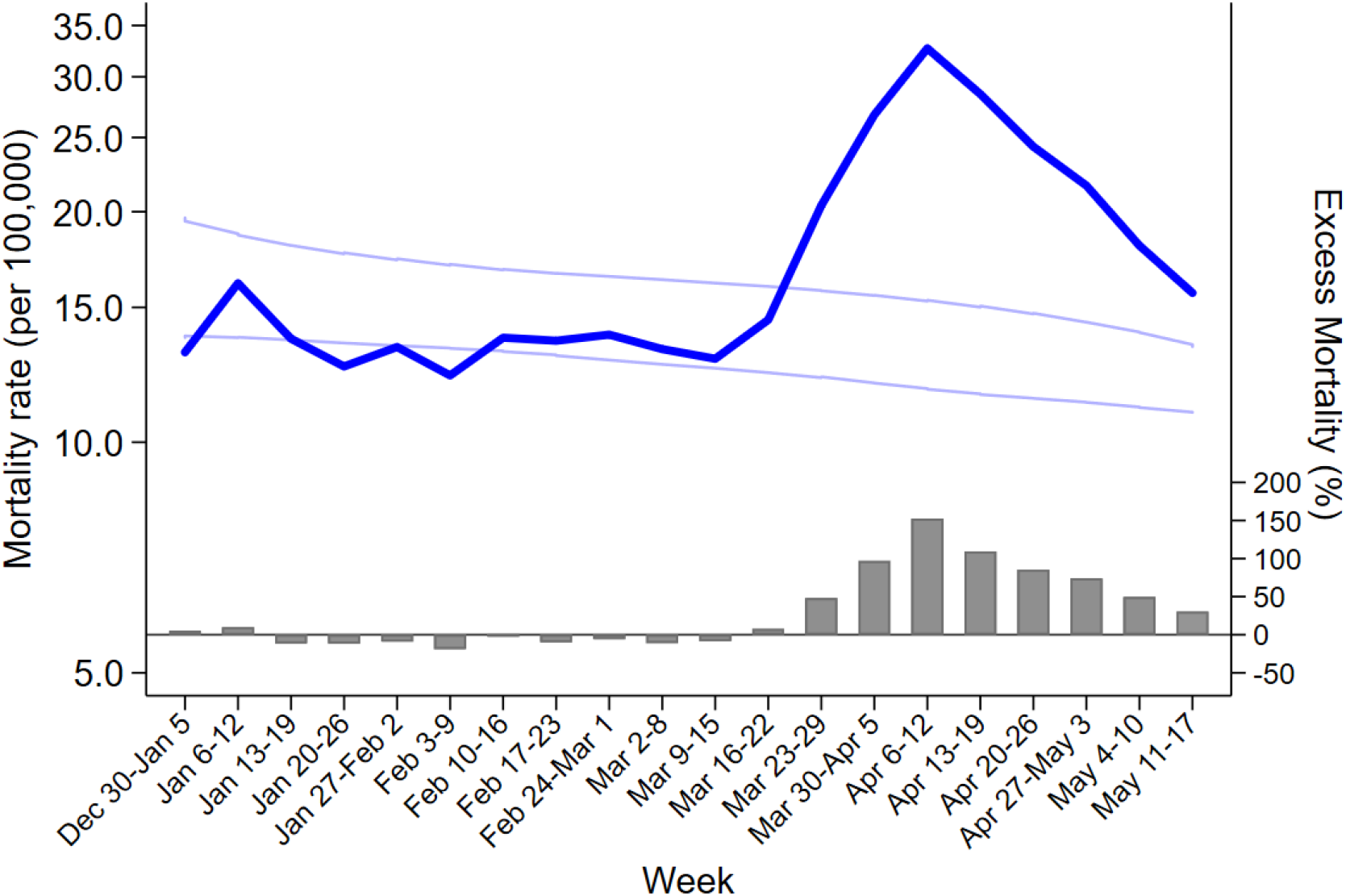
All-cause mortality rates (per 100,000 persons) in the Stockholm Region (N=2,379,792), Sweden, during the first 20 weeks of 2020 (dark blue line) and 95% confidence intervals for average mortality rates for the corresponding weeks during the five previous years (light blue lines). Bars depict the excess mortality during the first 20 weeks of 2020 in comparison to the average five-previous-years mortality.

During the outbreak, the highest excess mortality was recorded among DeSOs in the lowest tertiles for income, education level, share of Swedish-born and share of gainfully employed, with numbers reaching 171%, 162%, 178%, and 174%, respectively, for the peak week of 6-12 April 2020 (**Figure 2**). When further disaggregating socioeconomic tertiles according to the share of 70+ year-old people (dichotomized by the median), the highest excess mortality rates were observed among most deprived but also youngest DeSOs. The corresponding numbers for the peak mortality week (i.e. 6-12 April 2020) were 215% for the youngest DeSOs with lowest income, 221% for the youngest DeSOs with lowest education level, 198% for the youngest DeSOs with lowest share of Swedish-born, and 232% for the youngest DeSOs with lowest share of gainfully employed (**Figure 3**). Even within the most deprived DeSOs, there was a 1.2 to 1.7-fold increase in excess mortality between those with a lower vs. higher share of older people, depending on the socioeconomic indicator considered (week of i.e. 6-12 April 2020).

**Figure 2.**
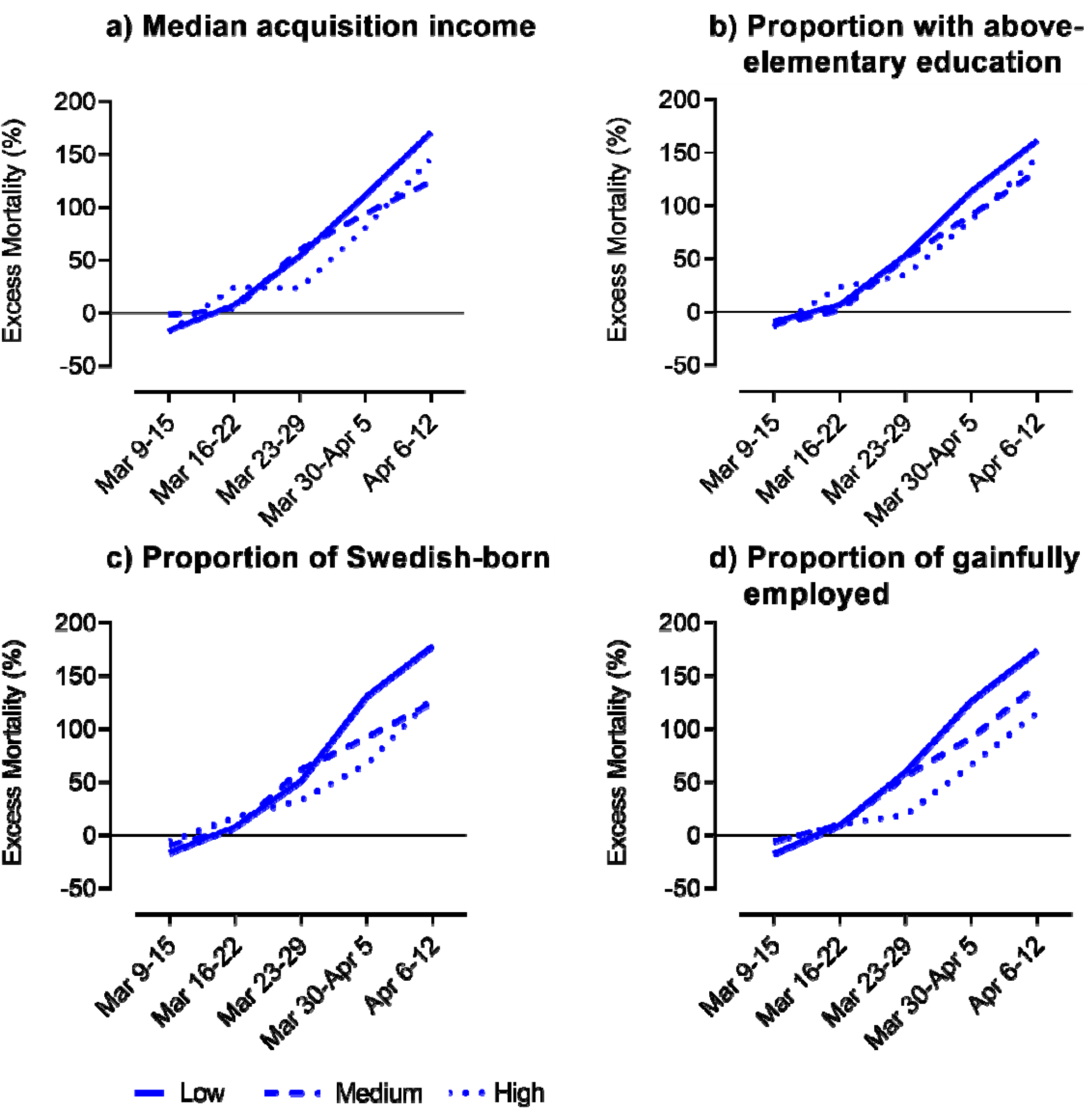
Average excess mortality across the 1,287 DeSOs of the Stockholm Region (N=2,379,792), Sweden, by levels (low, medium and high tertile) of socioeconomic indicators during the COVID-19 outbreak (i.e. March 9^th^ to April 12^th^ 2020). Note: Demographic Statistics Areas (DeSO) produced by Statistics Sweden (SCB) gather groups of around 1500 inhabitants (range: 700 and 2700) and are built within the municipal boundaries across Sweden. Excess mortality calculated comparing the weekly mortality rates during the weeks 11-15 of 2020 with the average mortality rate recorded for the corresponding weeks during the five previous years.

**Figure 3.**
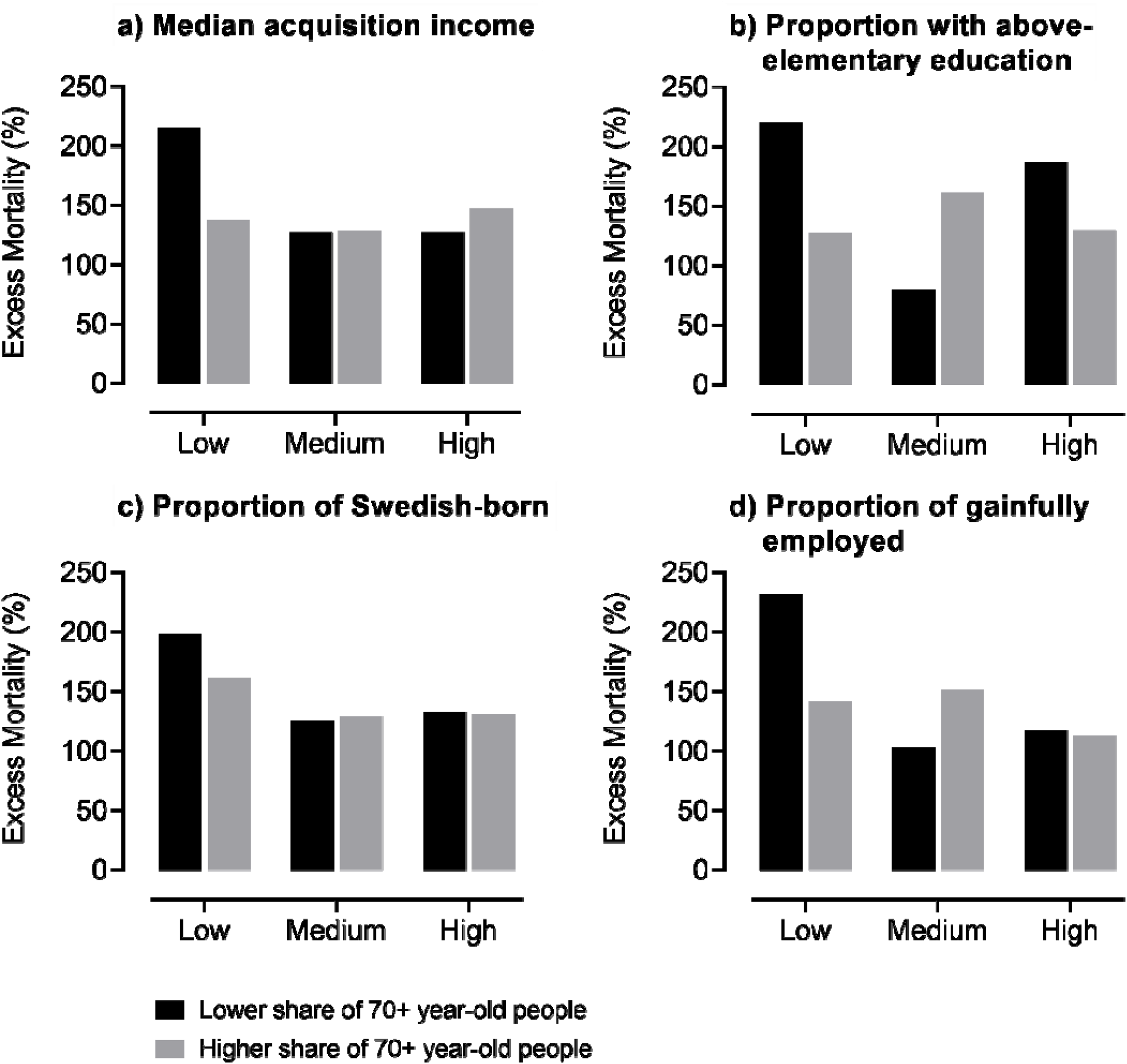
Average excess mortality across the 1,287 DeSOs of the Stockholm Region (N=2,379,792), Sweden, by levels (low, medium and high tertiles) of socioeconomic indicators and share of 70+ year-old people (below and above median) during the peak of the COVID-19 outbreak (i.e. week of April 6-12 2020). Note: Demographic Statistics Areas (DeSO) produced by Statistics Sweden (SCB) gather groups of around 1500 inhabitants (range: 700 and 2700) and are built within the municipal boundaries across Sweden. Excess mortality calculated comparing the weekly mortality rates during the week of April 6-12 2020 with the average mortality rate recorded for the corresponding week during the five previous years.

## Discussion

In the present work we found that in the Stockholm Region the negative impact of the COVID-19 outbreak was disproportionally born by older adults and people living in deprived areas with a higher proportion of young people, which underlines the prognostic role of the interplay between old age and a context of social vulnerability.

Our observation that the risk of severe COVID-19 is not uniformly distributed across population groups corresponds to other empirical evidence. Age is the main risk factor for COVID-19-related death, with over two thirds of Sweden’s deaths to date being in people aged 80+ years^2^ for whom, according to our results, an excess mortality of 193% was found. Age paces the progressive accumulation of multiple chronic conditions, which starts during adult life but exerts its full burden after the 60-70^th^ decade, when more than one in two individuals are affected by two or more chronic diseases (i.e. multimorbidity).^8^ Multimorbidity increases individuals’ risk of developing physical and cognitive impairments and facilitates the development of infectious diseases, such as pneumonia, due to viral and bacterial agents.^9–12^ Such lack of resilience to stressful events configures a condition of biological frailty, which is displayed by a considerable share of older adults, and has been recently identified as an independent risk factor for intensive care need and in-hospital death among older adults affected by COVID-19.^13,14^

Protracted social inequalities in non-communicable diseases and socially-patterned health determinants are being magnified by the COVID-19 pandemic, which places a disproportionate burden on those who are socioeconomically vulnerable.^15^ These individuals have higher rates of almost all known underlying clinical risk factors that increase the severity and mortality of COVID-19.^16^ Social determinants of health, governing where people work, live, and age, are the likely drivers of the social inequalities in COVID-19’s outcomes. In addition to influencing COVID-19’s chronic comorbidities, life-long socioeconomic adversity may lead to a suppressed immune response due to psychosocial stress increasing the likelihood of infection.^17^ Reduced incomes may lead to home overcrowding, which increases the risk of contagion too. Environments characterized by increased deprivation may have lower access to healthcare, even in universal healthcare systems, whereas reduced educational level and lower health literacy may impede access to and understanding of public health advice.^15^ Overall, rather than being socially neutral as claimed in the early days of the pandemic, COVID-19 exacerbates existing social inequalities in health and disease.

Along the lines of what other countries have suggested as an exit strategy from COVID-19 lockdown, Swedish policy has been directed to reduce intergenerational contact primarily among older people from the beginning. However, according to the idea of segmented shielding, almost as important as taking strict precautions to avoid infection among the most vulnerable (i.e. the older population), is extending such safety measures to their close regular contacts: those who live with them, the relatives who visit them and/or the social workers who care for them (i.e. the shielders).^6^ Whether Sweden’s less-restrictive approach to containing COVID-19 by promoting “social distancing” instead of compulsory quarantine has been more or less successful is yet to be examined in international comparative studies, but what seems to be evident from our study is that such strategy may have been less effective in deprived areas with high proportions of young people. Factors like occupational environments (e.g. service sector and other jobs requiring physical contact with others), an active social life, the shortage and lack of adapted preventive information, as well as overcrowded and intergenerational cohabitation may have increased the likelihood of the virus exposure among the so-called “shielders” living in these areas, with a direct impact on their vulnerable older populations, as shown by excess mortality rates surpassing 200% in these areas. Offering quarantine facilities to help people in crowded households to isolate themselves has been one important measure launched by some municipalities in the Stockholm Region, but perhaps not enough. The dissemination of translated guidelines to reduce the spread of COVID-19, for example, has claimed to be done with an unnecessary delay.^18^ Setting the maximum number of people allowed to gather in public events to 50^19^ might also have been insufficiently restrictive, given the propensity among younger people to meet in smaller groups.

Our findings have important implications for future shielding strategies, under the likely scenario of future resurgences of the COVID-19 infection^20^ -or other pandemics- and the widely acknowledged need to target interventions (e.g. confinement, social distancing, active surveillance) and direct healthcare resources towards the subgroups at increased risk of developing the severest forms of the disease.^21^ While different models of stratified shielding have already been suggested,^22,23^ these rarely account for contextual sociodemographic factors, which is likely to lead to insufficiently discriminatory public health responses given their lack of calibration not only to the people but also to the areas involved. In this regard, the real-time tracking of people’s individual characteristics and their sociodemographic context might ease the implementation of effective preventive and containment strategies. The lack of availability of electronic records with wide individual and contextual data coverage has in fact been pointed out as one of the weaknesses of the current response to the COVID-19 pandemic in developed countries, and referred to as one of the most important strategies to successfully cope with future similar scenarios.^24^

### Study limitations

For confidentiality reasons, we did not have data on DeSO-level mortality, which prevented us from obtaining variance estimates within DeSO groups. Given the lack of availability of individual-level data, we cannot ascertain the age of subjects contributing to the exceptionally high rates of excess mortality seen in the deprived young neighborhoods of the Stockholm Region. Still, we have reasons to believe that they are most likely 65 years or older, as is the case for the rest of the population. If, in the worst-case scenario, all people dying in these areas were below 65, the mortality rate for this age group during e.g. the peak week of 6-12 April 2020 would not make up for the actual number of deaths observed during that week (expected: 16 deaths; observed: 132-150 deaths depending on the socioeconomic indicator considered). Considering that the 2019-2020 winter in Stockholm was one of the mildest on record, the observed mortality in the period before the COVID-19 outbreak was in the low range in comparison with previous years.^25^ Thus, it is likely that the calculated excess mortality is underestimated. In fact, when comparing the mortality rate of the first week of April 2020 with that of three weeks earlier, we observed an excess mortality of 146%. On the other hand, it should be noted that the lower-than-average mortality preceding the outbreak, may have increased the “pool” of frail persons and thus contributed to the excess mortality during the outbreak.

## Conclusion

Future COVID-19 related strategies of social distancing, confinement, active surveillance, and eventually vaccination, will necessarily have to consider individuals’ risk of morbidity and mortality if they were to contract the virus, which implies accounting also for their sociodemographic context. In the Stockholm Region, for instance, vulnerable individuals living in deprived areas with a high proportion of young people will require fine-grained prevention strategies.

## Data Availability

Dara for this study were provided by Statistics Sweden, SCB (https://www.scb.se/en/)

## Acknowledgments

We thank Tomas Johansson, Ann-Sofie Davidsson and Christina Grandelius from Statistics Sweden for the assistance in the extraction of mortality and census data.

## Author’s contributions

ACL, DLV, TB and SD developed the study concept and design. ACL, DLV and DR performed the data analysis. ACL, DLV and SD drafted the manuscript. ACL, DLV, DM, TB, LF and SD interpreted the data. All authors provided critical revisions and approved the final version of the manuscript for submission.

